# Enhancing data integrity in Electronic Health Records: Review of methods for handling missing data

**DOI:** 10.1101/2024.05.13.24307268

**Authors:** Amin Vahdati, Sarah Cotterill, Antonia Marsden, Evangelos Kontopantelis

## Abstract

**Introduction:** Electronic Health Records (EHRs) are vital repositories of patient information for medical research, but the prevalence of missing data presents an obstacle to the validity and reliability of research. This study aimed to review and category ise methods for handling missing data in EHRs, to help researchers better understand and address the challenges related to missing data in EHRs.

**Materials and Methods:** This study employed scoping review methodology. Through systematic searches on EMBASE up to October 2023, including review articles and original studies, relevant literature was identified. After removing duplicates, titles and abstracts were screened against inclusion criteria, followed by full-text assessment. Additional manual searches and reference list screenings were conducted. Data extraction focused on imputation techniques, dataset characteristics, assumptions about missing data, and article types. Additionally, we explored the availability of code within widely used software applications.

**Results:** We reviewed 101 articles, with two exclusions as duplicates. Of the 99 remaining documents, 21 underwent full-text screening, with nine deemed eligible for data extraction. These articles introduced 31 imputation approaches classified into ten distinct methods, ranging from simple techniques like Complete Case Analysis to more complex methods like Multiple Imputation, Maximum Likelihood, and Expectation-Maximization algorithm. Additionally, machine learning methods were explored. The different imputation methods, present varying reliability. We identified a total of 32 packages across the four software platforms (R, Python, SAS, and Stata) for imputation methods. However, it’s significant that machine learning methods for imputation were not found in specific packages for SAS and Stata. Out of the 9 imputation methods we investigated, package implementations were available for 7 methods in all four software platforms.

**Conclusions:** Several methods to handle missing data in EHRs are available. These methods range in complexity and make different assumptions about the missing data mechanisms. Knowledge gaps remain, notably in handling non-monotone missing data patterns and implementing imputation methods in real-world healthcare settings under the Missing Not at Random assumption. Future research should prioritize refining and directly comparing existing methods.

## Introduction

Electronic Health Records (EHRs) have firmly established themselves as essential repositories of patient data, serving pivotal roles in medical statistics and diverse research objectives (1). These data offer a wealth of information, enabling researchers to investigate various health-related phenomena (2). However, one pressing challenge researchers face when utilising EHRs is the issue of missing data, which can introduce bias and affect the validity of their findings (3, 4). Real-world EHR data often have high levels of missingness, presenting a challenge for statistical analyses (5). Imputing missing data in EHRs is crucial for ensuring data integrity and facilitating reliable healthcare decisions (3, 4). When it comes to handling missing data in EHR, it is a critical aspect that can significantly enhance the precision of research and reduce bias. Missing data, if not addressed properly, can lead to skewed results and unreliable conclusions (3, 6). An Electronic Health Record (EHR) is characterized as a digital archive of patient information, accessible to authorized users across various healthcare settings, primarily aimed at facilitating comprehensive healthcare delivery (7). Data that was not initially gathered for research purposes often exhibits missing values, a common occurrence in EHR datasets. Consequently, effective management of missing data holds significant importance within the realm EHRs (8).

Missing data is commonly classified into three distinct types, determined by the underlying reasons for its absence. Missing completely at random (MCAR) is a term used to describe a situation where the occurrence of missing data is purely random and is independent of both observed and unobserved variables (9). Missing values in the specific variable don’t show any clear differences compared to the values that were observed (10). E.g. if a blood pressure is missing from a dataset, it could be because someone accidentally forgot to record it. In such cases, the randomness of the missing data implies that the calculations made using the available data should ideally be free from any bias (11). When data are MCAR, the resulting parameter estimates are ideally entirely free from bias. However, a significant limitation of conducting an analysis solely on complete cases, which essentially overlooks the missing data issue, is the resultant loss of statistical power. This loss is not negligible in many cases, particularly in multiple regression analyses with a high number of predictors. Even minimal missing data in individual variables can lead to excluding a significant portion of the dataset from the study [14].

Shifting focus to the missing at random (MAR) category, here, the absence of data is influenced by the information that is available and recorded in the dataset (12). Unlike MCAR, in MAR, there is a connection between what we see and what’s missing. However, this link is only with the observed data, not with the missing values themselves (13). For example, in a large dataset, there is missingness regarding blood pressure data. Upon further analysis, it was found that other variables systematically differ between observed and unobserved values of blood pressure, particularly when considering cardiovascular disease and age. Patients without cardiovascular disease and younger patients had more missing blood pressure data compared to older patients with cardiovascular disease (11). The term ‘informative missingness’ is sometimes preferred over MAR, and its presence can be evaluated using logistic regression to explore the relationship between predictor values and outcome missingness [14].

In the most complex scenario, when data is missing not at random (MNAR), the variable’s unobserved value is associated with its absence’s underlying cause. This unobserved value can serve as a predictor or, more concerning, as an outcome. In this case, producing valid results is challenging due to the absence of straightforward methods. One approach is to conduct multiple sensitivity analyses to examine the impact of missing data on the outcomes of the study (14). One example is how Body Mass Index (BMI) data is recorded. BMI is more often collected for people who are overweight or obese than for those who are not, because health professionals are aware of the link between weight and various health conditions. Sometimes, when patients are not obese, their records might be left blank, and it is hard to tell if the data is missing or if the person just isn’t obese. Figuring out this kind of missing data, called Missing Not at Random (MNAR), is tough because it’s hard to know why the data is missing (15, 16).

Despite the significance of this methodological concern, there is a noticeable gap in the literature regarding a comprehensive examination of the various approaches and strategies for handling missing data in EHR-based observational studies. While there are existing reviews on this topic, they often focus on narrower aspects or specific methods (16–18). Therefore, there is a compelling need for a scoping review that can offer a broad overview of the methodological approaches used in addressing missing data within the context of EHR-based observational studies.

## Methods

This analysis was conducted using the scoping review methodology defined by Arksey and O’Malley (19). A scoping review approach is beneficial for identifying existing peer-reviewed research and pinpointing key evidence. Scoping reviews are designed to chart the evidence landscape rather than deliver critically evaluated and synthesised findings, eliminating the need for quality assessment of the included studies. Literature published and accessible in full text on EMBASE by October 2023, focusing on missing data imputation in EHRs and presented either as a review article or through a novel approach, was considered eligible for inclusion.

We employed a customised systematic search strategy that combined keywords pertinent to missing data imputation and electronic health records (Table. 1) (20, 21). Eligible for inclusion were both review articles and original studies proposing approaches to managing missing data in EHRs.

**Table 1.**
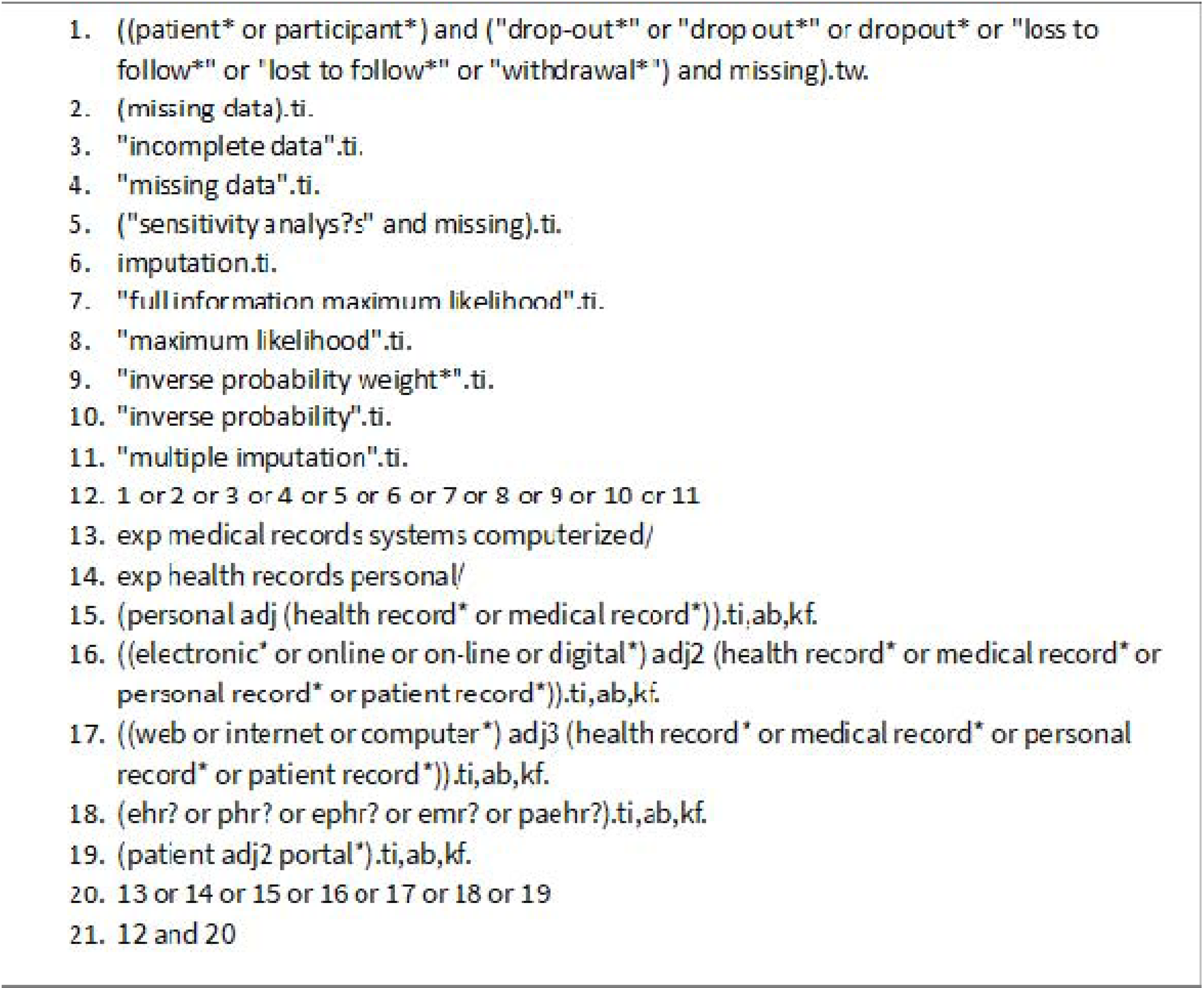
A search strategy was developed by combining relevant keywords related to imputing missing data and electronic health records, utilizing Boolean operators.

After removing duplicates, we began by systematically reviewing titles and abstracts against the inclusion criteria to exclude any that were clearly ineligible, followed by screening the remaining full texts to identify relevant papers. We also searched the publication lists of the final pool of included papers for relevant references. Data extraction focused on imputation techniques, the characteristics of datasets, assumptions about missing data and the type of article.

## Results

A total of 101 articles were found, and two of them were excluded as duplicates, as indicated in the PRISMA scoping review flowchart (Figure. 1). The remaining 99 documents were screened by title and abstract. 21 studies were selected for full-text screening. Five studies were eligible for data extraction. Four articles were also found through a manual search, which brings the total number of studies that will be screened for full text to nine.

**Figure 1.**
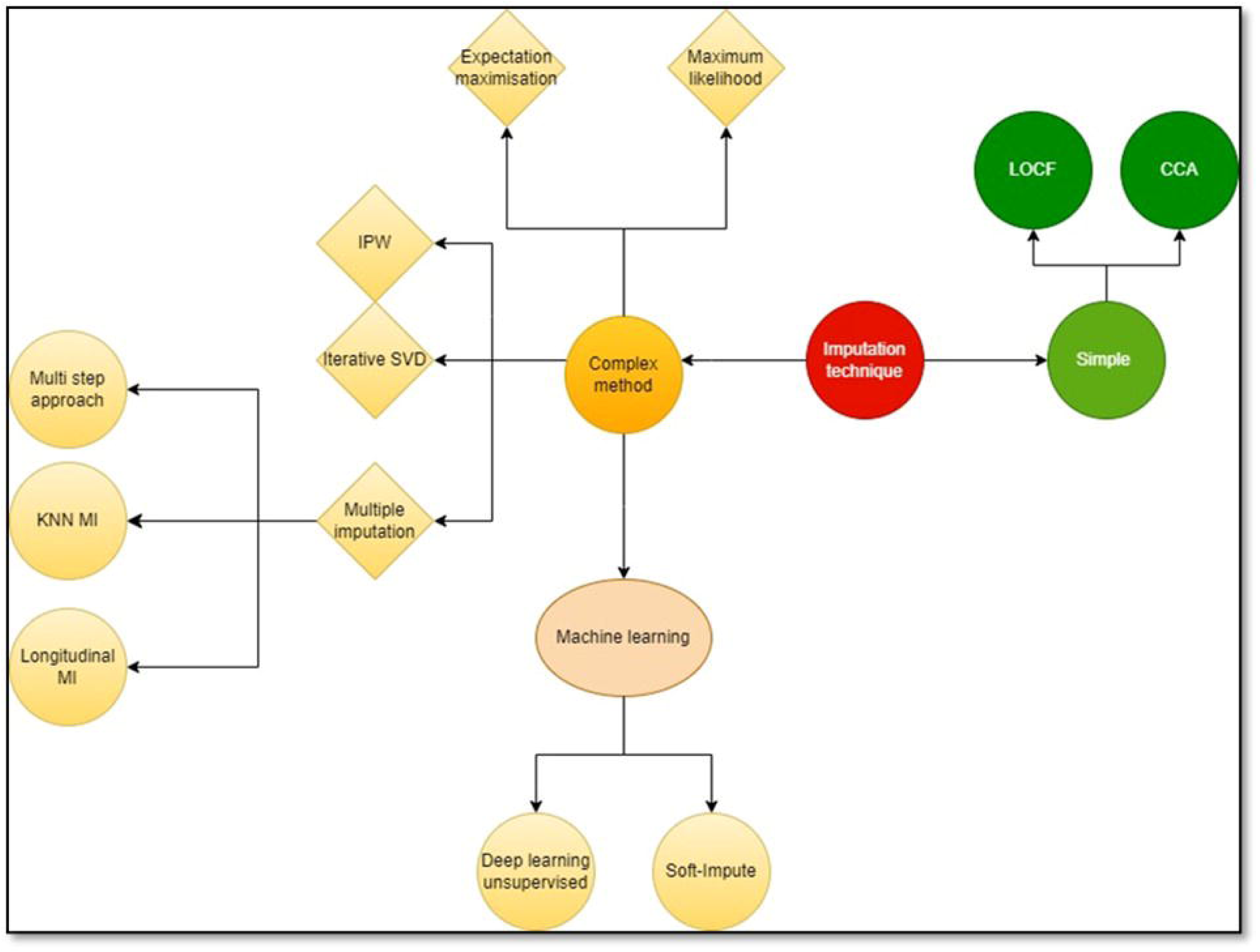
Prisma flowchart of literature search.

Within the nine articles, thirty-one imputation approaches were identified, classified into 10 distinct imputation methods, each applied under specific assumptions, using simple or more complex imputation methods (Figure. 2).

**Figure 2.**
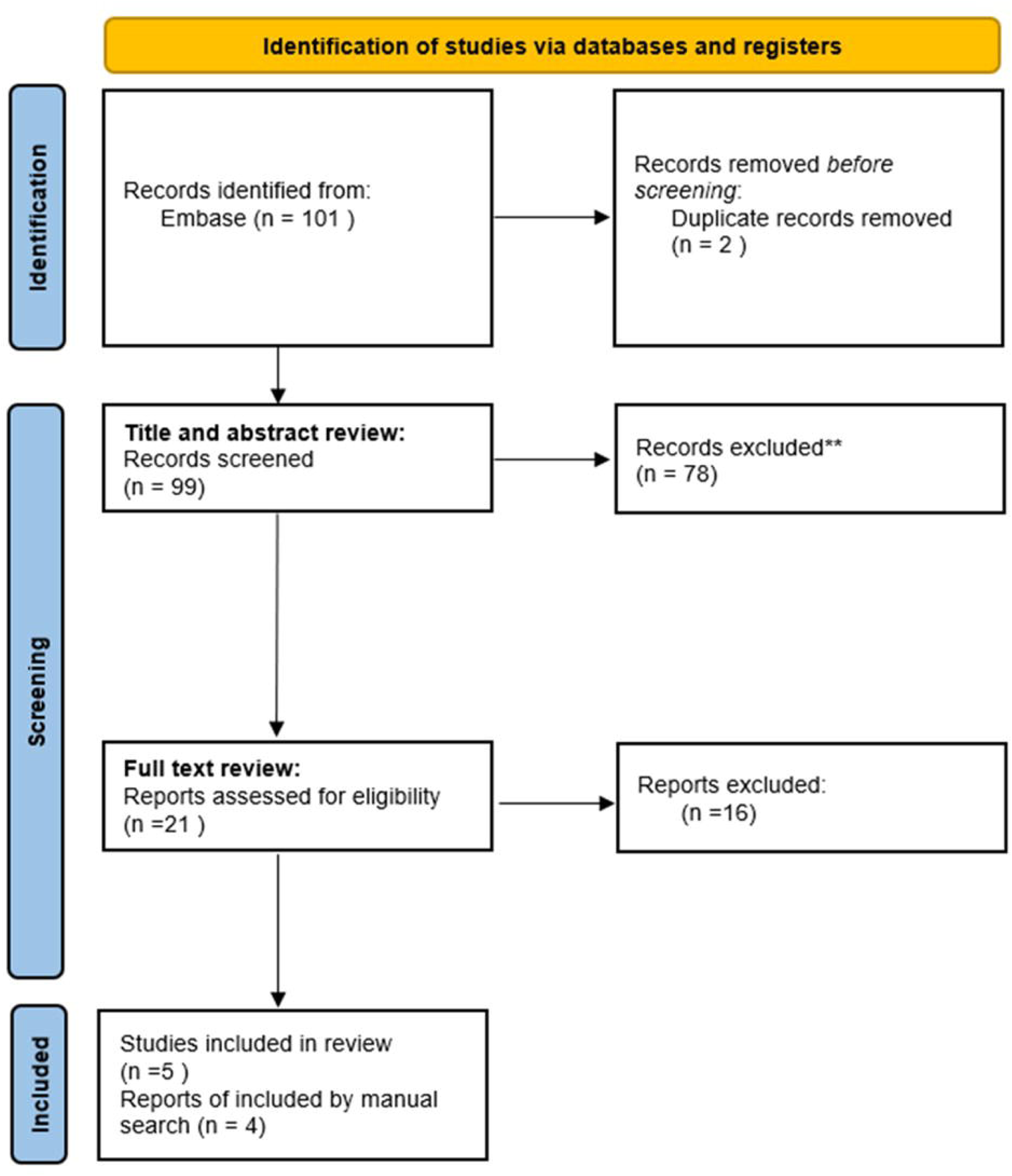
Imputation methods.

The screened articles encompassed various contributions to the field: three papers provided reviews of existing imputation methods, two engaged in simulation studies, two introduced novel methodologies, one established a general framework for imputing missing data, and one conducted a comparative analysis of different autoencoder methods.

Certain studies proposed imputation methods suitable for a broad spectrum of EHRs (16, 22, 23), while others focused on specific data types such as time series (17), marginal structural models (18), and longitudinal datasets (15, 24, 25). Additionally, a unique study employed a simulation study for comparing different imputation methods (26). The following synthesis provides a comprehensive overview of the diverse landscape of imputation methodologies applied to address missing data in EHRs (Table 2).

**Table 2.**
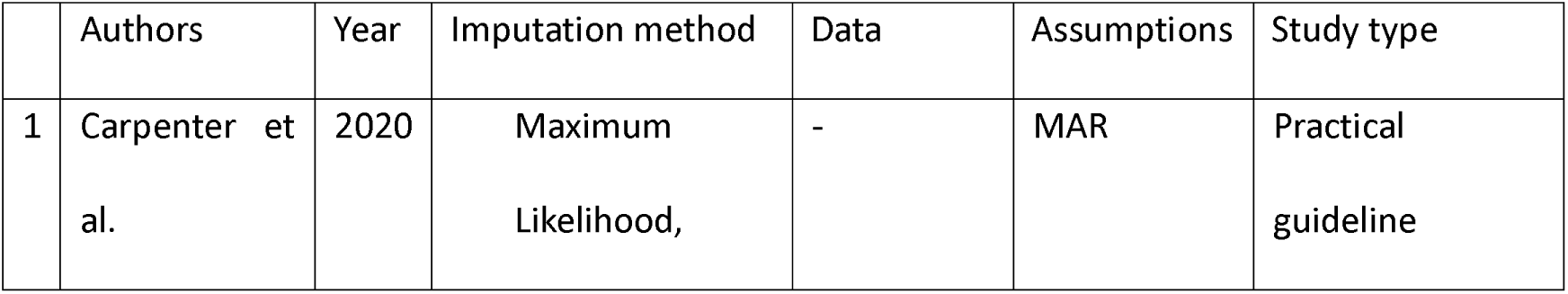
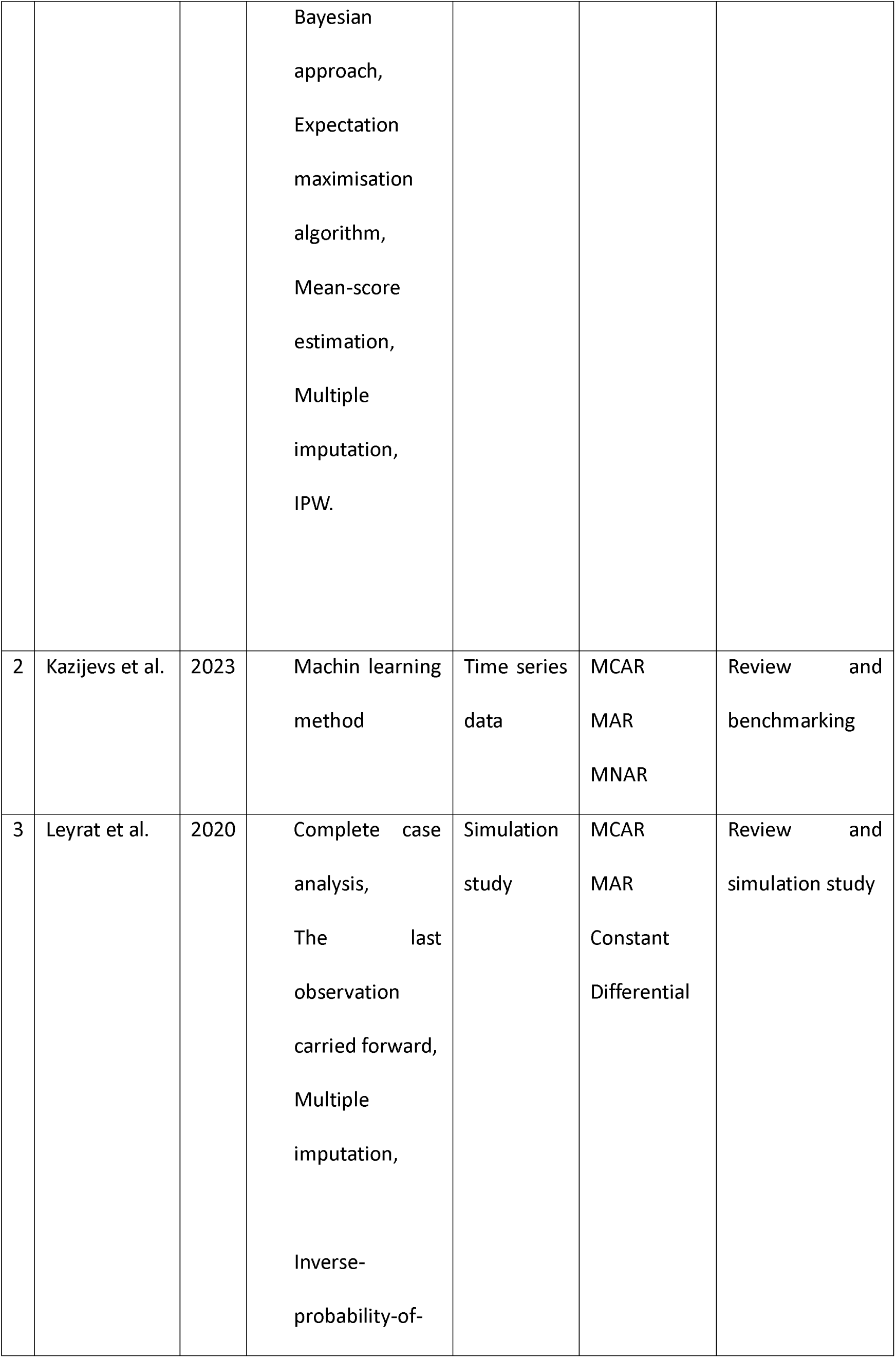
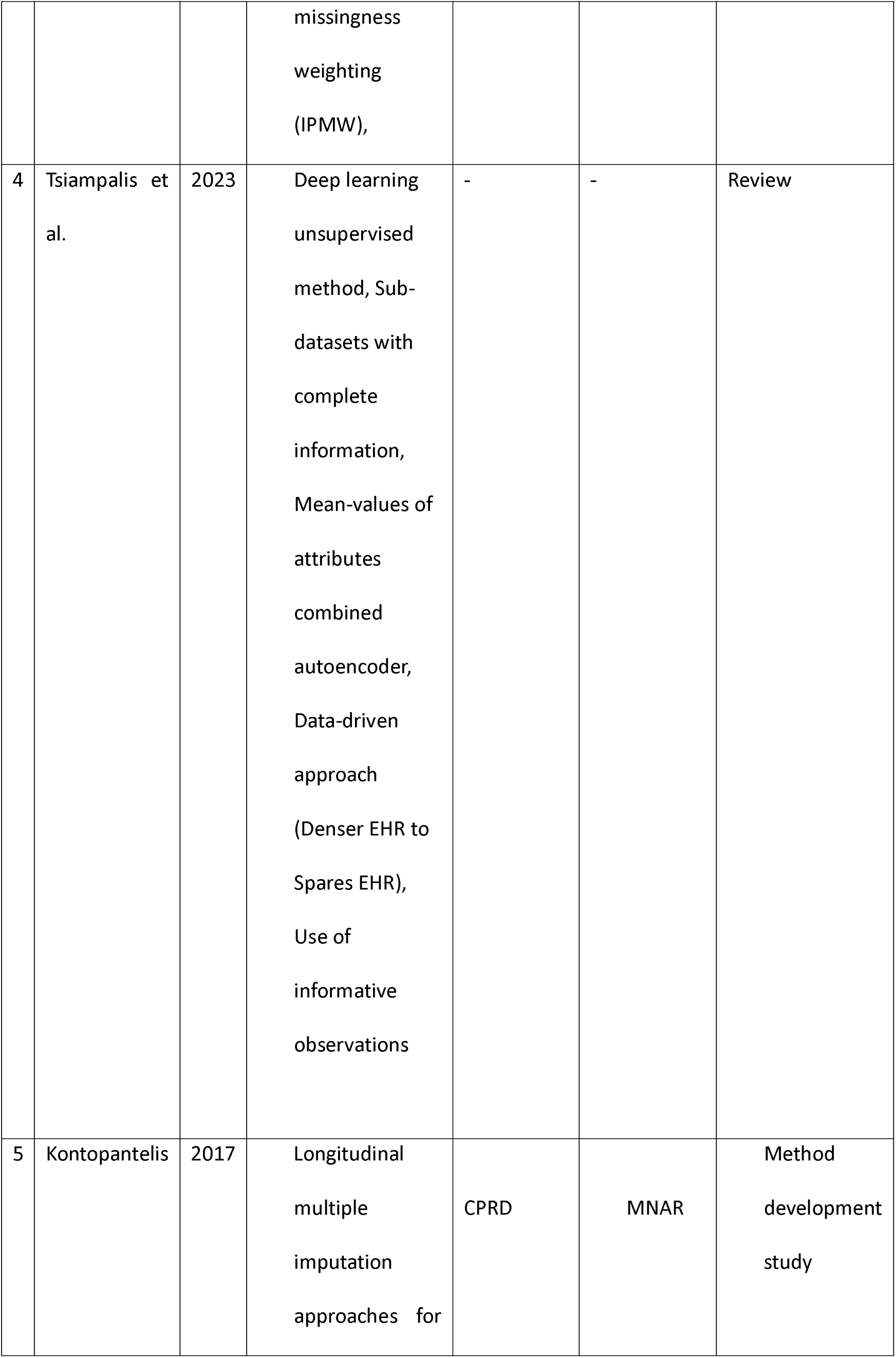
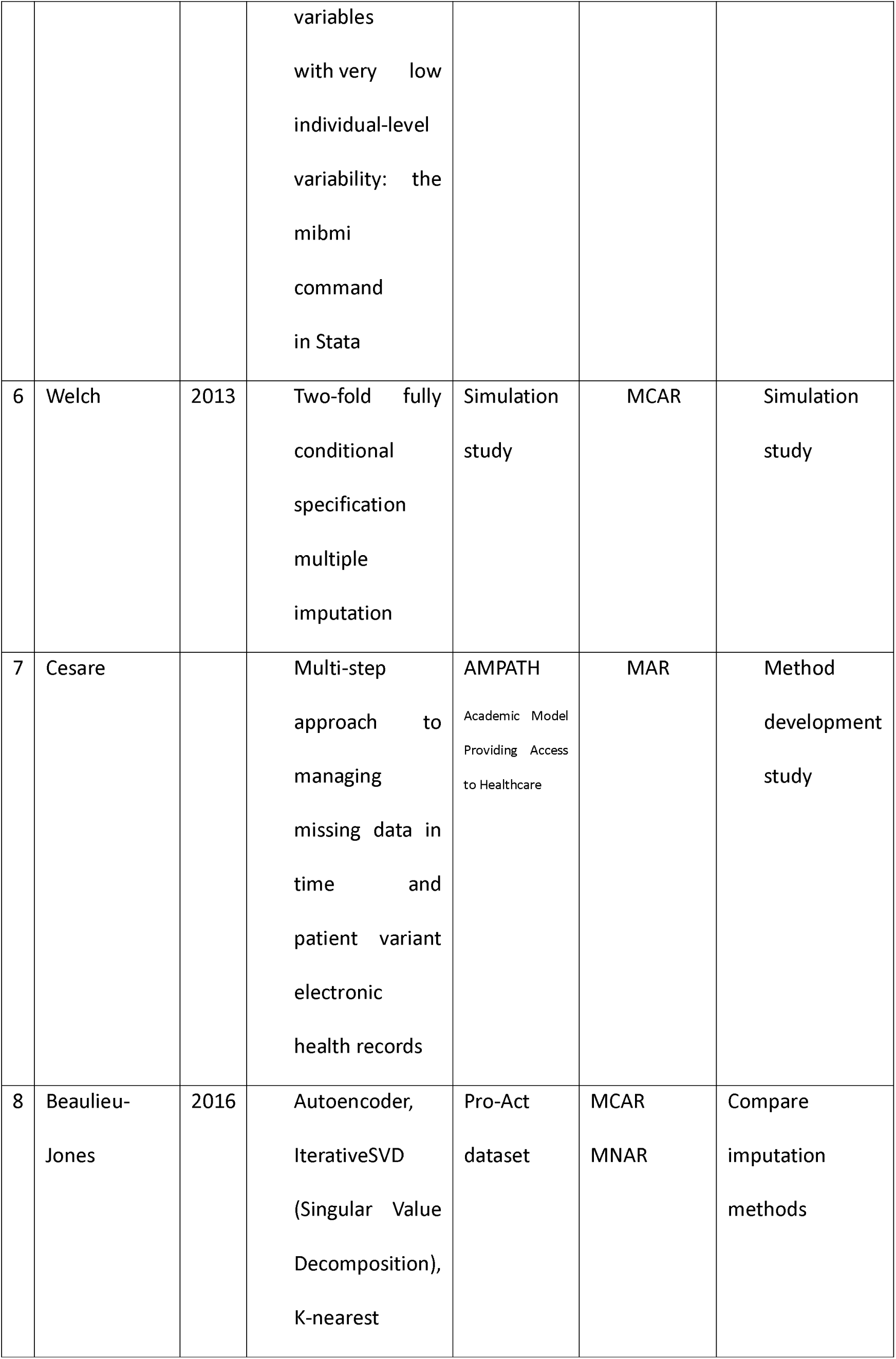
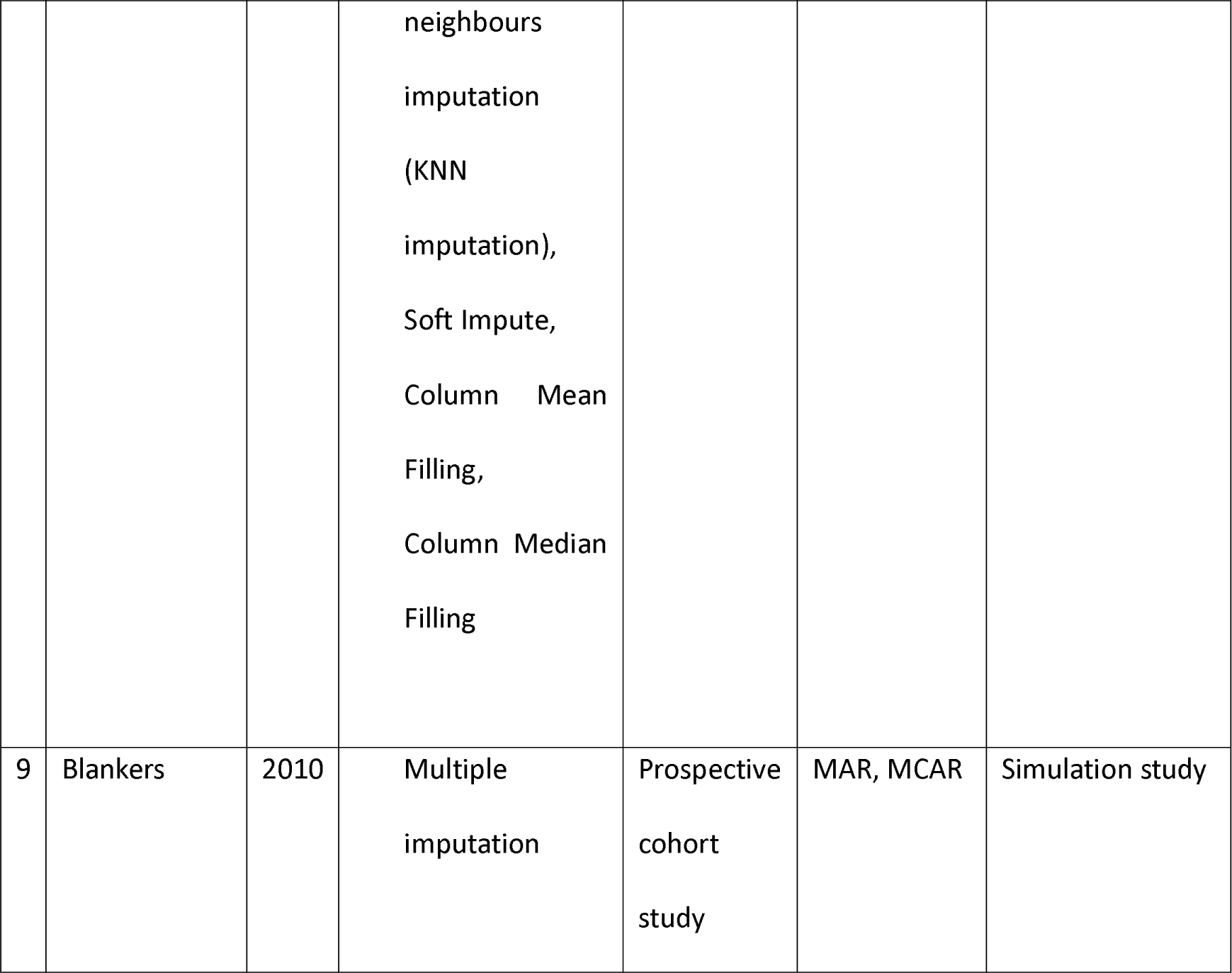
Characteristics of Included Studies.

|  | Authors | Year | Imputation method | Data | Assumptions | Study type |
| --- | --- | --- | --- | --- | --- | --- |
| 1 | Carpenter et al. | 2020 | Maximum Likelihood, | - | MAR | Practical guideline |
|  |  |  | Bayesian approach, Expectation maximisation algorithm, Mean-score estimation, Multiple imputation, IPW. |  |  |  |
| 2 | Kazijevs et al. | 2023 | Machin learning method | Time series data | MCAR<br>MAR<br>MNAR | Review and benchmarking |
| 3 | Leyrat et al. | 2020 | Complete case analysis,<br>The last observation carried forward,<br>Multiple imputation,<br>Inverse-probability-of- missingness<br>weighting<br>(IPMW), | Simulation study | MCAR<br>MAR<br>Constant<br>Differential | Review and simulation study |
| 4 | Tsiampalis et al. | 2023 | Deep learning unsupervised method, Sub-datasets with complete information, Mean-values of attributes combined autoencoder, Data-driven approach (Denser EHR to Spares EHR), Use of informative observations | - | - | Review |
| 5 | Kontopantelis | 2017 | Longitudinal multiple imputation approaches for variables<br>with very low<br>individual-level<br>variability: the<br>mibmi<br>command<br>in Stata | CPRD | MNAR | Method development study |
| 6 | Welch | 2013 | Two-fold fully<br>conditional<br>specification<br>multiple<br>imputation | Simulation<br>study | MCAR | Simulation<br>study |
| 7 | Cesare |  | Multi-step<br>approach to<br>managing<br>missing data in<br>time and<br>patient variant<br>electronic<br>health records | AMPATH<br>Academic Model<br>Providing Access<br>to Healthcare | MAR | Method<br>development<br>study |
| 8 | Beaulieu-Jones | 2016 | Autoencoder,<br>IterativeSVD<br>(Singular Value<br>Decomposition),<br>K-nearest neighbours<br>imputation<br>(KNN<br>imputation),<br>Soft Impute,<br>Column Mean<br>Filling,<br>Column Median<br>Filling | Pro-Act<br>dataset | MCAR<br>MNAR | Compare<br>imputation<br>methods |
| 9 | Blankers | 2010 | Multiple<br>imputation | Prospective<br>cohort<br>study | MAR, MCAR | Simulation study |

A combined count of 32 software packages dedicated to imputation methods was discovered across the four statistical software: R, Python, SAS, and Stata. Notably, machine learning techniques for imputation were absent from devoted packages in SAS and Stata. Among the 9 imputation methods searched, implementations were accessible for 7 methods across all four software platforms (Table 3).

**Table 3.**
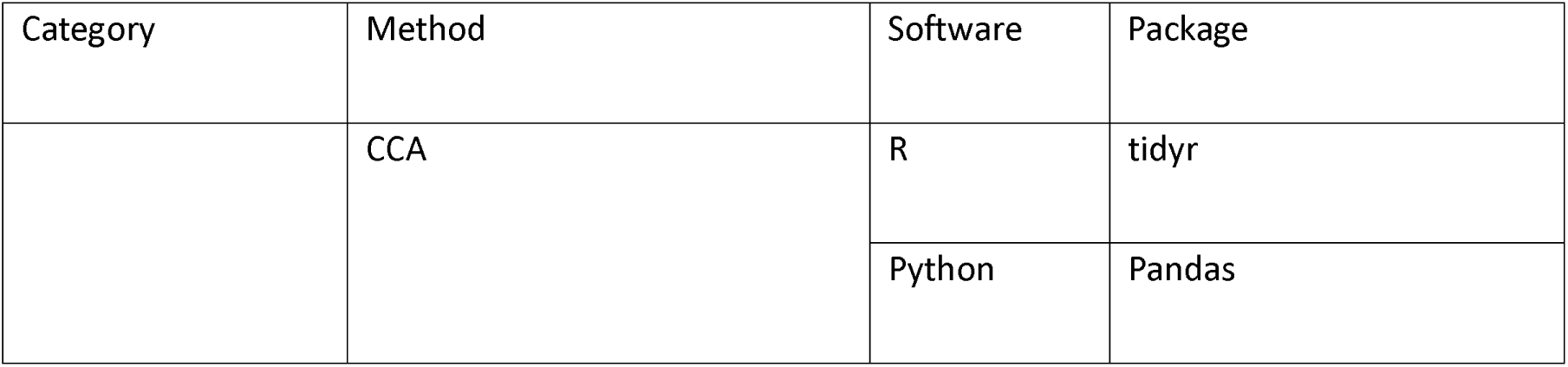
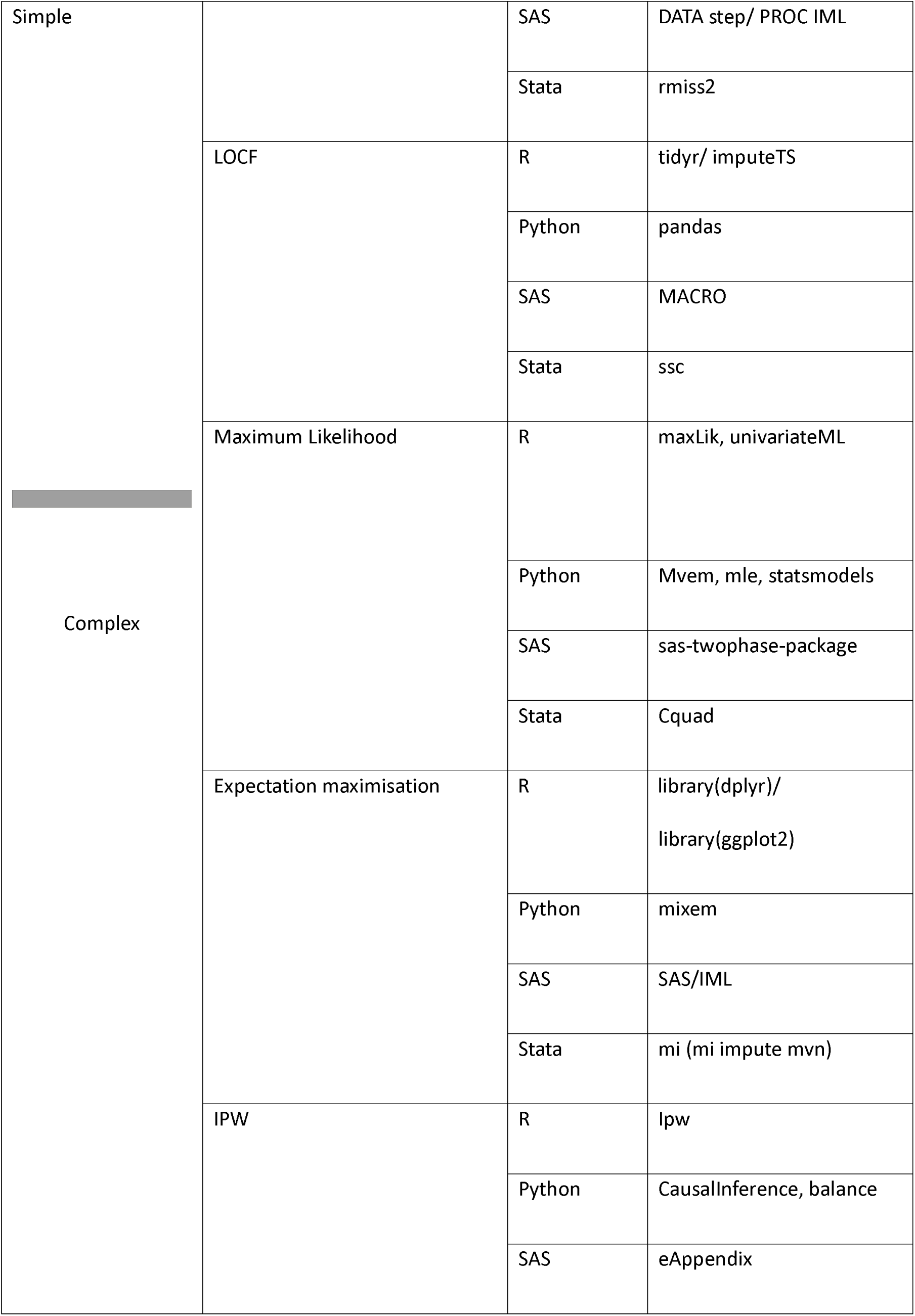
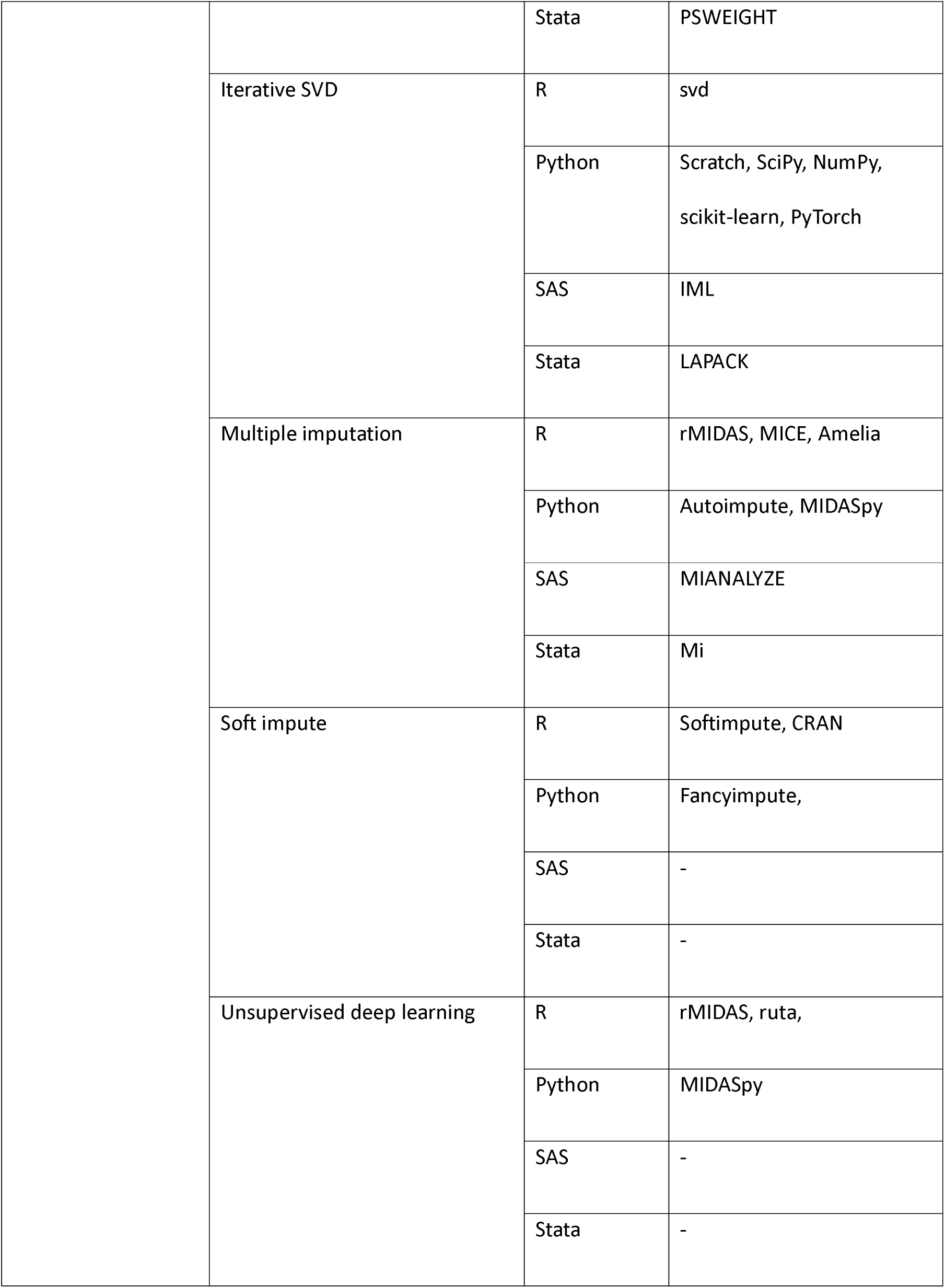
Imputation methods package.

## A. Simple methods

### A.1 Complete case analysis

Complete-case analysis (CCA), or listwise deletion, exclusively employs the data records that keep no missing values for any pertinent variable required for analysis, hence excluding observations that have a missing value for any of the variables included in the analysis (27). CCA can produce estimates of effect size that are biased, inefficient and/or underpowered. These issues become more pronounced when there is a high level of missing data, with loss of power and also higher levels of bias if the data missingness mechanism is MAR or MNAR. (28). The fundamental assumption underlying CCA is that the data are MCAR.

### A.2. Last observation carried forward (LOCF)

The LOCF method is a historically popular statistical tool in longitudinal studies involving repeated measurements (29). In this approach, when a value is missing from a later visit, it is filled in with the previously recorded value of the participant. This leads to an analysis where through a simple form of imputation, the affected variables can be regarded as complete (18). However, a critical shortcoming of LOCF is its inclination to provide inaccurate estimates for missing values, notably when the dependent variable is on an upward (or downward) trajectory over time. LOCF is prone to bias, potentially leading to either an underestimation or overestimation of the actual effects of the treatment. The method’s suitability is questionable even in situations where the missing data is purely random (30).

## B. Complex imputation methods

### B.1. Multiple Imputation (MI)

Multiple imputation is a commonly used method that replaces missing values by creating plausible numbers based on the distributions and connections of observed variables in the dataset (31). MI utilises this technique to estimate the missing values, resulting in multiple datasets considered ‘full’. We use the observed data to evaluate the distribution of the partially observed variables, given the fully known variables (32). Subsequently, we employ this estimation to fill in the missing data. The rationale behind using multiple imputations is that the imputed data can never possess the identical characteristics as the observed data. Instead, they are generated from the predicted distribution of the missing data, given the observed data under the assumption of MAR. This process operates under a specific Bayes model that accurately represents both the observed data and the mechanism responsible for missing data (32).

Two steps are involved in multiple imputations: 1) Creating replacement values, or “imputations,” for missing data and repeatedly doing so to develop numerous data sets with the missing information replaced. Multiple Imputation (MI) fills in the missing values using statistical properties of the data, such as the relationships and distributions of variables in the dataset. 2) Analysing and integrating the many imputed data sets (33, 34). Following the execution of the planned statistical analysis (such as regression or t-test) on each imputed data set individually (stage 2), the desired estimates (e.g., the average difference in outcome between a treatment group and a control group) from all the imputed data sets are merged into a single estimate using standard combining methods. The benefit of using MI in statistical analysis is that it may handle problems other than traditional missing data problems. MI is a widely used method for managing missing data, and it is offered in many software programmes (35, 36).

Further refinement in data imputation is achieved using Multiple Imputation by Chained Equations (MICE, also called Fully Conditional Specification (FCS)), a sophisticated technique crucial for datasets with complex variable relationships (37). MICE works by creating multiple dataset copies, filling missing values with placeholders, and then employing regression models to predict these values (38). The pooled predictions from these multiple imputed datasets lead to the selection of final values, effectively handling both categorical and continuous data. Even after employing MICE, some values, especially dates, might be inaccurate. The subsequent step involves selecting representative values for each patient to minimise outlier impacts and manually adjusting date variables to ensure accuracy (25). The primary limitations of this approach involve the time-invariant nature of the MICE imputation model and the lack of consideration for spatial autocorrelation among nearby clinics.

A Bayesian multiple imputation method is introduced to handle left-censored multivariate data commonly found in environmental and biomedical research (39). This approach addresses the difficulty of assigning explicit values for observations below the limit of detection (LOD), especially in longitudinal data settings.

Also, k-nearest neighbours (KNN) can be used with bootstrap and sequential imputation to get multiply imputed data (40). The idea behind utilising KNN for missing values is that a point value may be estimated by the values of the points nearest to it, depending on other factors. For categorical variables, the mode of the nearest neighbours is utilised, whereas for numeric data, the mean of the nearest neighbours is employed. The k-nearest neighbours algorithm requires searching through all complete cases and selecting the k examples most relevant to a particular missing information (Nearest neighbour selection for iteratively KNN imputation). The KNN imputation technique encounters two significant hurdles: firstly, determining the optimal value of k in advance, and secondly, selecting the most appropriate k nearest neighbours [53].

### B.2. Maximum likelihood

Maximum likelihood imputation is a theoretically robust approach for estimating parameters in regression models when dealing with missing data (41). This technique includes estimating a set of parameters that maximise the chance of obtaining the observed data (42). The method is characterised by the explicit articulation of the likelihood of the intended data, integration over missing values to ascertain the likelihood of observed data, and the subsequent maximisation of this likelihood to derive maximum likelihood estimates. This methodological rigour ensures a principled and statistically sound treatment of missing data, especially in complex and unbalanced study designs (22, 43). A limitation of maximum likelihood methods is the need for relatively large data sets (44).

Maximum likelihood and MI procedures have been methodically developed with a foundational assumption of MAR (45). Note that if the missingness mechanism strays from the MAR assumption, as observed in MNAR scenarios, methods such as maximum likelihood and multiple imputation might produce biased results, especially in small longitudinal samples exhibiting non-normality (43). In instances of varying population distributions, Maximum Likelihood is preferred over MI when the distribution is non-normal (46).

### B.3. Expectation-Maximisation (EM) algorithm

Expectation maximisation offers an iterative approach to maximising the likelihood estimation by using latent variables (47). This algorithm is designed to maximise the likelihood of observed data by iteratively refining parameter estimates. The EM algorithm consists of two distinct phases: Expectation and Maximization. In the Expectation phase, the algorithm begins by imposing a value for the missing or latent variables based on the current estimates of the parameters. This step involves calculating the expected values of the missing data given the observed data and the current parameter estimates. Essentially, it estimates the posterior distribution of the missing variables. This imputation process is crucial for establishing a foundation for subsequent parameter refinement. Following the Expectation step, the Maximization phase comes into play. In this step, the algorithm evaluates the parameters that maximise the expected log-likelihood obtained from the first step. It involves adjusting the parameter values to enhance the fit between the observed and imputed data. The Maximisation step essentially serves as a parameter update based on the newly imputed values, optimising the likelihood function. The iterative nature of the EM algorithm involves repeating these two steps until a convergence criterion is met, indicating that the algorithm has reached a stable solution. Convergence is often achieved when there is minimal change in the parameter estimates between successive iterations. One notable advantage of the EM algorithm is its ability to provide consistent estimates of means and covariance matrices, which are essential for characterising the underlying distribution of the data. However, EM may be computationally intensive and requires a sufficiently large sample size to ensure the reliability of the parameter estimates (48–50).

### B.4. Inverse probability weighting (IPW)

In this method, complete cases are weighted by the inverse of their probability of being complete cases. Table four illustrates a basic idea. Because the likelihood of missing data being dependent on its value introduces bias, the observed mean is 13/6 instead of the whole dataset’s mean of 2 (i.e., the actual values of the nine observations). The third row of table four displays the estimated chance of observing the data when considering the group variable and assuming MAR given the group. This assumption was validated by cross-referencing with the entire data (22).

**Table 4.** Example for inverse probability weighting imputation technique.

| Group | A |  |  | B |  |  | C |  |  |
| --- | --- | --- | --- | --- | --- | --- | --- | --- | --- |
| Full data | 1 | 1 | 1 | 2 | 2 | 2 | 3 | 3 | 3 |
| Observed data | 1 | ? | ? | 2 | 2 | 2 | ? | 3 | 3 |
| Probability of observation given group | 1/3 | - | - | 1 | 1 | 1 | - | 2/3 | 2/3 |

The weights, which are the inverses of the observation probabilities, are used to compute a weighted mean using this estimate. Therefore, to make up for the three observations in group A, we give each observation a weight of 3. Next, we compute the weighted mean:

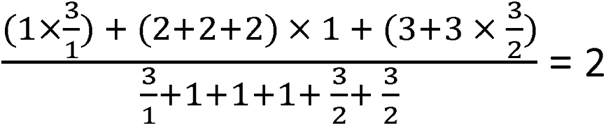

Since the IPW analysis relies on estimated weights from observed data, it only applies under MAR (51). Statistical analysis is complicated by IPW. To begin, it considers only complete records when re-weighting them, which results in the elimination of any data point that is lacking values. As a result, the study may not be as comprehensive or representative as it could have been because data from these missing variables cannot be recovered (52). If covariates are not MAR given the dependent variable, a complete records analysis might be more suitable, necessitating the inclusion of the dependent variable in the weight model. It can be challenging to estimate weights precisely when relevant variables in the weight model have missing values, further complicating the inclusion of these variables. Lastly, IPW results can be affected by big weights and the weight model chosen, which can make it hard to choose. Finding a suitable weight model becomes much more of a challenge when there is no explicit documentation on handling these problems (22).

There is a modified method of IPW called Inverse Probability of Missingness Weighting (IPMW). In studies with time-varying confounding and data gaps, IPMW is used. While effective in monotone missing data scenarios, like participant dropout, IPMW becomes complex with non-monotone missing patterns, where data intermittently lacks in various variables. This complexity increases with multiple time points and incomplete variables. Additionally, the broader inverse probability weighting (IPW) method, which combines weights for each missing variable, can lead to variable weights and less accurate treatment effect estimates. “IPW” refers to the general approach, with “IPMW” denoting its specific uses for missing data (53). IPMW is used in Marginal Structural Model (MSM) for dealing with missing confounder in non-randomised longitudinal studies (18). Marginal structural models (MSMs) are commonly used to estimate causal intervention effects in longitudinal nonrandomized studies (54).

### B.5. Iterative SVD (Singular Value Decomposition)

To understand Iterative SVD, it is essential first to grasp the concept of standard SVD. Singular Value Decomposition is a matrix factorisation technique used in many fields, including signal processing, statistics, and machine learning (55). SVD is a mathematical technique that decomposes a matrix into its constituent elements, providing insight into its structure. It decomposes a matrix Ā into three other matrices U, Σ and V^T^ where U and Σ and V^T^ are orthogonal matrices, and Σ is a diagonal matrix containing singular values. This decomposition can capture the underlying structure of the

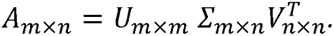

Iterative Singular Value Decomposition (SVD) addresses missing values in the initial data matrix. Given that SVD necessitates an entirely populated matrix, an initial imputation is performed by substituting missing values with mean or median values or utilising row/column averages. After the initial imputation, standard SVD is performed on the modified data matrix. This yields the matrices U and Σ and V^T^, providing a lower-dimensional representation of the data. The missing values are subsequently recalculated utilising the knowledge acquired from the SVD decomposition. This entails utilising the singular value decomposition (SVD) components to restore the data matrix and replacing the previously imputed values with new estimations. The final two steps are performed iteratively. During each iteration, the Singular Value Decomposition (SVD) is performed on the matrix that has been updated with imputed values. Subsequently, these values are adjusted depending on the new decomposition (56, 57).

Singular Value Decomposition (SVD) offers notable advantages, including significantly higher accuracy compared to simple row averages across diverse datasets (58). It excels in analysing time-series data with low noise levels, effectively estimating gene expression based on temporal regulation patterns (57). However, SVD has limitations, requiring complete matrices for operation. Imputation strategies, such as substituting row averages for missing values, are necessary. Additionally, SVD’s performance is sensitive to data type, exhibiting potential challenges in non-time series datasets lacking clear expression patterns (59). Its linear regression nature in lower-dimensional space may result in diminished performance for non-time series data, where expression patterns are often less distinct (57). Despite these considerations, SVD remains a powerful tool, particularly suited for specific data characteristics and applications.

### B.6. Longitudinal imputation approach

The two-fold fully conditional specification (FCS) technique is an adaptation of FCS as multiple imputation technique, that takes account of the temporal structure of longitudinal data (60, 61). The method commences by selecting a time window width, for instance, 1-, 2-, or 3-time blocks, establishing the range of time blocks around the target time point for imputation. During this stage, the typical FCS imputation technique is utilised to fill in missing values for each variable at a particular time (61). This is done by employing regression models incorporating values from adjacent time points within the designated time window (61, 62). This method enables the inclusion of temporal dependencies within the imputation model. Once the within-time imputation for a time block is finished, the same process is done for all time blocks in the dataset. This step guarantees that the imputation model considers the longitudinal evolution of the data (61, 62).

The two-fold FCS method improves the collection of temporal correlations in longitudinal EHR data by considering adjacent time blocks throughout the imputation process. This simplifies the process of imputation models and minimises the likelihood of collinearity and overfitting (61). The system enables the utilisation of several modelling approaches tailored to specific variable types, thus accommodating the wide range of data types commonly encountered in EHRs. The two-fold technique can result in more accurate estimates compared to traditional FCS/MICE when there are longitudinal and time-dependent patterns present, reducing bias.

Like other multiple imputation approaches, the two-fold FCS functions assume that data are MAR. The algorithm can be computationally intensive, especially with large datasets and comprehensive time windows (24). Bespoke longitudinal imputation algorithms, like the `mibmi` code in STATA, are customized for specific parameters such as BMI over time. These algorithms start with outlier detection and employ standard and regression-based cleaning methods for BMI computation. While effective in generating multiple imputed datasets, they come with drawbacks including high computational costs, exclusion of patients with limited BMI records, and the necessity for careful extrapolation to avoid inaccuracies.

## C. Machine learning methods

Machine learning methods for imputing missing data operate by dividing datasets into training and test sets and learning from observed variables. There are several methods, using supervised, semi-supervised, or unsupervised machine learning techniques [59]. Like other imputation methods, machine learning methods do not work with all kinds of data [60]. Their performance could change depending on the value type, which include numeric, non-numeric, text, graph, etc. So, it is essential to fully understand the underlying data patterns before doing data imputation to ensure that the best machine learning method is chosen for handling missing data situations correctly and effectively [61]. Selecting the proper machine learning imputation method depends on the dataset and analysis aim [61]. When compared to standard imputation methods, the use of machine learning for imputing missing values has shown improved performance in prediction and data analysis [62]. Non machine learning techniques may result in a smaller sample size and more bias since they limit variability [63]. Nevertheless, due to recent technological progress, machine learning capitalises on substantial computational resources to tackle these obstacles efficiently through precise estimation of absent values, thus enhancing the performance of data analysis [64]. Current method proposals that use machine learning techniques can have their crucial improvements in accuracy, performance, and time consumption brought to light through research and analysis [24]. Different machine learning methods have been used in EHRs for imputing missing data in the screened articles. We will introduce them more.

### C.1. Deep learning unsupervised method

Deep learning unsupervised is an imputation technique specifically tailored to handle longitudinal patient data, defined by the progression of the disease over time (63). This strategy primarily involves collecting data by grouping essential clinical variables that an expert physician has determined. The data, characterised by missing values denoted as “NA,” is structured into records associated with patients via unique identifiers. The methodology entails an initial preprocessing stage in which the data is converted into a matrix structure. The matrix has n rows (records) and m columns (variables) and is subjected to z-score normalisation for numeric variables and one-hot encoding for categorical variables. The approach’s critical element resides in utilising a deep autoencoder framework. This system consists of two distinct encoders: Encoder1, which is dedicated to creating embeddings at the record level, and Encoder2, which captures the heterogeneity at the patient level. These encoders operate by projecting the input into a hidden space, and then a decoder combines these embeddings and processes them through numerous layers that include nonlinear transformations. The parameters are adjusted in the last stage of the process, and the patient data is re-entered to provide comprehensive datasets. The utilisation of a deep learning architecture in this method enables the modelling of intricate inter-variable interactions, which is crucial for effectively filling in missing values in cardiovascular patient data. The strategy effectively minimises imputation mistakes and biases in cardiovascular patient care by utilising patient-level heterogeneity and temporal patterns, representing a significant development in data processing (64). The deep autoencoder framework, with multiple layers and non-linear transformations, can be computationally intensive (65). Training and fine-tuning such models may require significant computing resources and time, which could be a practical limitation in certain healthcare settings. Deep learning models, particularly intricate ones such as autoencoders, frequently suffer from a lack of transparency and interpretability (66, 67). The comprehension of the model’s process in determining imputed values or the rationale behind its conclusions might be difficult, which can cause apprehension in clinical environments where interpretability is essential.

### C.2. Soft-Impute

This technique is especially relevant in fields such as machine learning and data science, where dealing with incomplete data is a common challenge. Soft-Impute is grounded in spectral regularisation, using convex relaxation techniques to fill in missing values in large matrices. The core of this method lies in its use of the nuclear norm as a regularise. The nuclear norm, essentially the sum of the singular values of a matrix, helps maintain the low-rank structure of the solution and avoids overfitting, which is crucial when dealing with large datasets. The process of Soft-Impute is iterative. It employs a soft-threshold Singular Value Decomposition (SVD) in each iteration to update the missing elements of the matrix (68). Through iterative updates using SVD, Soft-Impute efficiently replaces missing values, gradually improving the matrix’s completeness. One of the critical strengths of Soft-Impute is its ability to compute a regularisation path, offering a series of solutions corresponding to different values of the regularisation parameter. This capability allows the method to find the optimal balance between the complexity of the model and it’s fit to the data, making it highly effective for practical applications. In terms of scalability and efficiency, Soft-Impute stands out. With high computational efficiency, it can handle huge matrices, such as those encountered in the Netflix challenge for predicting user movie ratings. This efficiency stems from its fast computation of a low-rank SVD of a dense matrix. Furthermore, the method has shown impressive performance, achieving good training and test errors compared to other state-of-the-art techniques [70].

## Discussion

The comprehensive review conducted in this study provides a valuable summary of imputation methods for addressing missing data in EHRs. With the increasing digitization of healthcare data, the issue of missing data has become a significant concern, as it can impact the reliability and validity of analyses derived from such data. In this study, we identified a total of 101 articles and chose nine studies for data extraction, which described 31 imputation approaches. These methods were divided into two primary groups: simple imputation methods and complex imputation methods. Simple methods included CCA and LOCF, while complex methods comprised MI, Maximum Likelihood, EM algorithm, IPW, Iterative SVD, longitudinal imputation approach, deep learning unsupervised method, and Soft-Impute. Researchers should carefully consider the characteristics of their data, the assumptions of each method, and the specific context of their study when selecting an imputation method. The choice of imputation method should align with the goals of the analysis and the nature of the missing data mechanism.

The performance of different imputation methods has been assessed in various studies. A comparison of eight imputation methods under different missing data mechanisms (MI, CCA, mean imputation, LOCF, HOT deck imputation, regression imputation, KNN, EM algorithm) suggested that MI exhibits the least standard errors compared to other methods, indicating its effectiveness in handling missing data (69). Simulation studies offer valuable insights, but it is crucial to validate these findings with real-world datasets to determine the practical applicability and reliability of multiple imputation (MI) in comparison to other imputation methods.

Comparison of multiple imputation methods, including chained equations, random forests, and denoising autoencoders, revealed distinct patterns across different missing data mechanisms. Chained equations and random forests showed reduced bias and comparable standard errors under MCAR. However, denoising autoencoders exhibited elevated bias in cases of MAR. Furthermore, all methods demonstrated increased bias proportional to the extent of missing data under MNAR conditions (70).

Assessing the performance of FCS and Two-Fold FCS methods in managing missing variables within longitudinal studies, particularly in estimating regression coefficients via a linear regression model, reveals that Two-fold FCS methods yield estimates that are slightly more biased and less precise compared to FCS (71, 72). Two-Fold FCS was specially adapted from FCS to deal with missing data in longitudinal studies, so it seems surprising that FCS would perform better than two-fold FCS in longitudinal data. This observed bias likely stems from the inherent limitation of these approaches in restricting variables within the univariate imputation models, consequently risking the omission of crucial information of the missing data (71). One study suggests that, in longitudinal data, imputation of data from patients with a similar pattern of data may outperform traditional MI methods (73). Multiple imputation (MI) is an effective method for dealing with bias caused by missing data in longitudinal studies, resulting in accurate parameter estimates.

A comparison of eight common statistical and machine learning imputation methods (simple imputation, regression, EM, MICE, KNN, clustering imputation, random forest, and decision tree) showed that KNN and RF were the most effective imputation methods in the cohort study dataset under the MAR assumption (74). Machine learning methods attempt to explore the relationship between variables and predict missing data more precisely. This superior performance of machine learning methods suggests their potential for handling complex data patterns and nonlinear relationships, thereby offering more accurate imputations in challenging datasets.

Understanding the assumptions behind each imputation method is essential for choosing the best one based on the dataset and missing data mechanism. Also, one should evaluate the computational complexity and resource needs when choosing imputation algorithms. Deep learning unsupervised methods can handle complex data patterns better but take a lot of processing power and time to train and fine-tune models. Although simpler algorithms like CCA and LOCF are more computationally efficient, they can be biased and inefficient. Existing methods address monotone missing patterns, temporal correlations in longitudinal data, and complex model interpretability, but they can be improved. The software available to researchers may influence their choice of method as not every method is available in each software.

Regarding knowledge gaps, firstly there is a lack of consensus regarding the superiority of certain imputation methods over others, particularly in scenarios involving complex missing data patterns. This discrepancy underscores the need for further comparative studies to evaluate the strengths and limitations of different imputation techniques comprehensively. Secondly, existing imputation methods may not adequately address the challenges posed by non-monotone missing data patterns, where missingness occurs sporadically or irregularly across the dataset. While methods like MI and machine learning algorithms have been proposed for handling monotone missing data, there is limited research on techniques specifically designed for non-monotone missingness. Developing robust imputation strategies tailored to these complex missing data patterns is crucial for improving the accuracy and reliability of analyses conducted on longitudinal datasets. Furthermore, there is a need for more research on the practical implementation of imputation methods in real-world healthcare settings. Many existing studies focus on theoretical comparisons of imputation techniques using simulated datasets or controlled experiments. While current approaches such as MI and other techniques have shown promise in dealing with missing data, further research is required to build viable solutions suited particularly to MNAR circumstances.

Imputation strategies for addressing missing data are often underutilized due to a prevalent practice of not explicitly reporting missing data (75, 76). This reporting failure adds to the knowledge gap on the kind and amount of missing data in epidemiological studies. Because of this, chances to use imputation techniques efficiently to solve problems with missing data are often missed or underutilized. The absence of imputation procedures in this study might potentially undermine the validity and reliability of the results. This highlights the need of thorough reporting and the use of suitable approaches to address missing data in research projects.

## Conclusion

Addressing missing data is an important aspect of the analysis of EHRs and various imputation methods are available to researchers. These methods range from simple to complex, each relying on assumptions that have been discussed in this paper.

When selecting imputation methods, key considerations include understanding the assumptions underlying each method, evaluating computational complexity, and effectively addressing monotone missing data patterns. Further research is needed to establish method superiority, especially in complex missing data scenarios, and to develop robust strategies for real-world healthcare applications.

## Data Availability

All data produced in the present work are contained in the manuscript.

